# Fibromyalgia, Chronic Fatigue Syndrome, and Multiple Sclerosis are associated with lower translocator protein binding affinity genotypes

**DOI:** 10.64898/2026.10.19.26365953

**Authors:** Chloe Jones, Jarred Younger

## Abstract

**Introduction:** Neuroinflammation is a known pathophysiologic mechanism in multiple sclerosis and a proposed mechanism in fibromyalgia and myalgic encephalomyelitis/chronic fatigue syndrome (ME/CFS). Positron Emission Tomography (PET) imaging of the translocator protein (TSPO) has been widely used to measure neuroinflammation, though second-generation ligands are highly sensitive to a single nucleotide polymorphism, rs6971, resulting in high, mixed, and low affinity binders. It is unknown whether this polymorphism is associated with neuroinflammatory disorders.

**Materials and Methods:** Eighty-six total participant genotypes for TSPO binding affinity were identified through Sanger sequencing (30 fibromyalgia, 21 ME/CFS, 11 multiple sclerosis, and 24 healthy controls). Associations between patient group and genotype were assessed with Fisher’s Exact test. Associations between genotype and symptom severity from self-report questionnaires were assessed with Kruskal-Wallis tests.

**Results:** In the present study individuals with fibromyalgia, (ME/CFS), or multiple sclerosis were more likely to have a mixed- or low-affinity genotypes compared to healthy controls. Presence of the low or mixed genotypes were also associated with greater self-reported fatigue severity and depressive symptoms.

**Discussion:** These findings should be confirmed in larger samples but suggest functional consequences of rs6971 polymorphism that may be related to the pathophysiology of pain and fatigue illnesses.

## Introduction

Myalgic encephalomyelitis/chronic fatigue syndrome (ME/CFS) and fibromyalgia (FM) are debilitating disorders characterized by chronic pain, fatigue, and cognitive symptoms, affecting approximately 1.3% [1] and 2% (Centers for Disease Control and Prevention, May 25, 2022) of the US population, respectively. Despite their high prevalence and clinical burden, their underlying etiopathologies remain poorly understood. Neuroinflammation has been proposed as a primary mechanism in both illnesses. In an inflamed state, microglia, the resident immune cells of the brain, release a cascade of proinflammatory cytokines, which can induce symptoms of pain, fatigue, and cognitive dysfunction [2]. Additionally, known neuroinflammatory diseases, such as multiple sclerosis, have overlapping symptomatology with FM and ME/CFS [3, 4]. The most direct evidence of neuroinflammation in ME/CFS and FM to date has been through the use of translocator protein (TSPO) positron emission tomography (PET) imaging [5-9].

TSPO-PET is one of the most widely used methodologies to assess neuroinflammation. Proinflammatory microglial activation in the brain is associated with upregulation of the mitochondrial TSPO 18 kDA. PET imaging of TSPO is therefore used to detect neuroinflammation in-vivo across a range of conditions, including glioblastoma [10], HIV [11], epilepsy [12], Alzheimer’s disease [13], multiple sclerosis [14] and in experimentally induced inflammation [15].

Contemporary TSPO PET ligands, including [^18^F]DPA-714, [^18^F]DPA-713, and [11C]PBR28, have good signal-to-noise ratios and TSPO specificity, but exhibit significant interindividual binding variability [16]. This variability is the result of a single nucleotide polymorphism (SNP), rs6971, on the TSPO gene [17]. This SNP results in a substitution from alanine (Ala) to threonine (Thr) (A147T) on the fifth transmembrane domain of TSPO. This codominant gene leads to three genotypes, C/C (Ala/Ala), C/T (Ala/Thr), and T/T (Thr/Thr). Those homozygous for the dominant allele (C/C) are high affinity binders for second-generation TSPO ligands. Those with one copy of the polymorphism (mixed affinity binders, C/T) express both high and low affinity binding sites. Those with two copies of the polymorphism (T/T) are low affinity binders and show negligible detectable signal. Therefore, low affinity binders are often excluded from TSPO PET studies, and the mixed or high affinity genotypes of individuals is identified to adjust for the significant impact on signal [18].

While collecting data for a TSPO-PET imaging study in ME/CFS, FM, and MS, we observed that patients more often had mixed or low affinity genotypes compared to healthy controls. We hypothesized that patients would be statistically more likely to present with the T/T or C/T genotypes when contrasted to healthy controls. We further hypothesized that lower TSPO affinity binding would be associated with greater fatigue, pain, and depressive symptom severity. Such a finding may indicate a functional consequence of the Ala to Thr substitution in the pathophysiology of pain and fatigue disorders.

## Materials and Methods

### Participants

Women with ME/CFS, FM, MS and healthy women between the ages of 18-65 in the Birmingham, Alabama area were recruited via online postings, flyers, and from our existing laboratory database to participate in a [^18^F]DPA-714 PET imaging study. Participants were enrolled between 2020 and 2025. The neuroimaging results of the FM patients compared to the HC have been previously published [7].

### Eligibility

Individuals were ineligible if they reported MRI contraindications, pregnancy or breastfeeding, a history of HIV, hepatitis C, cancer, blood disorder, autoimmune disorder (other than MS for the MS group), recent illness or infection (within 1 month of participation), or participation in experimental treatments. All participants who provided informed consent and provided blood for DNA genotyping were included in the present analysis. Only participants who completed further screening requirements were eligible for PET imaging [7].

FM participants met the 2016 American College of Rheumatology (ACR) case criteria [19] and reported daily pain of at least 6 out of 10. ME/CFS participants met Fukuda et al. criteria [20]. Only women with relapsing-remitting MS confirmed by physician diagnosis were included in the MS group. Healthy participants reported daily pain of 1 or lower and daily fatigue of 2 or lower on 0 to 10 scales.

### Protocol

The study protocol was approved by the Institutional Review Board at the University of Alabama at Birmingham (UAB) and conforms to the provisions of the Declaration of Helsinki. Individuals first underwent a phone screening to determine initial eligibility. All participants in the present study provided informed consent. Potential participants were then invited to attend an in-person consent and screening at the UAB Advanced Imaging Facility. Following informed consent, participants underwent blood-draw, urine pregnancy test, and completed several self-report questionnaires. Individuals with FM also underwent tender point examination with manual algometer and were excluded if fewer than 11 tender points were demonstrated according to ACR criteria.

### Genotyping

Venous blood was collected into two 4mL EDTA-coated tubes and prepared for processing at the UAB Specimen and Processing Biorepository Unit (SPAN). DNA was isolated manually using the Qiagen Puregene method. Optical density of 260/280 was determined using the Trinean Dropsense. The PCR product was directly sequenced by Sanger Sequencing. Participants were sequenced only for the TSPO gene.

### Questionnaires

Participants completed the Brief Pain Inventory [21], DePaul Symptom Questionnaire [22], and the Hospital Anxiety and Depression Scale [23]. Participants also self-reported demographic information.

### Analysis

The relationship between patient group (ME/CFS, FM, and HC) and rs6971 genotype (C/C (Ala/Ala), C/T (Ala/Thr), or T/T (Thr/Thr)) was analyzed using Fisher’s exact test.

Whether the presence of the rs6971 polymorphism (C/T or T/T) was associated with symptoms was assessed with Wilcoxon Two-Sample tests due to the ordinal nature of self-reported symptom scores. Symptoms of interest included self-reported fatigue ratings on the DePaul Symptom Questionnaire (0–100), average pain severity scores from the Brief Pain Inventory (0–10), and total anxiety and depression scores from the Hospital Anxiety and Depression scale (0–21). Symptom scores were compared between all three genotypes with Kruskal-Wallis tests and significant group differences were followed up with Steel-Dwass pairwise comparisons. Genotypes were also compared on the factor of race/ethnicity with Fisher’s exact test. All statistical analyses were performed with statistical software JMP version 18.2.2 (Cary, NC).

## Results

### Sample characteristics

A total of 86 participants were included in the present analyses: 30 FM, 21 ME/CFS, 11 MS, and 24 HC. All participants were female with a mean age of 42.44 (SD = 12.89). The sample was 71.43% Caucasian/White (n = 60), 23.81% African-American/Black (n = 20), and 4.76% Asian (n = 4). Two participants did not report their race/ethnicity. There were 48 C/C, 30 C/T, and 8 T/T genotypes in the sample.

### Genotype

The results of Fisher’s exact test indicated a significant association between genotype and patient group (*p* = 0.049). Participants with ME/CFS, FM, and MS were more likely to exhibit the rs6971 SNP compared to healthy controls (Fig 1). Among the healthy controls, 79.17% were high-affinity binders, compared to 42.86–48.39% across the patient groups. When analyzed separately, all patient groups showed significant differences in genotype compared to HC; FM (*p* = 0.030), ME/CFS (*p* = 0.045), and MS (*p* = 0.027). There was no association in this sample between genotype and race/ethnicity (*p* = 0.87).

**Fig 1.**
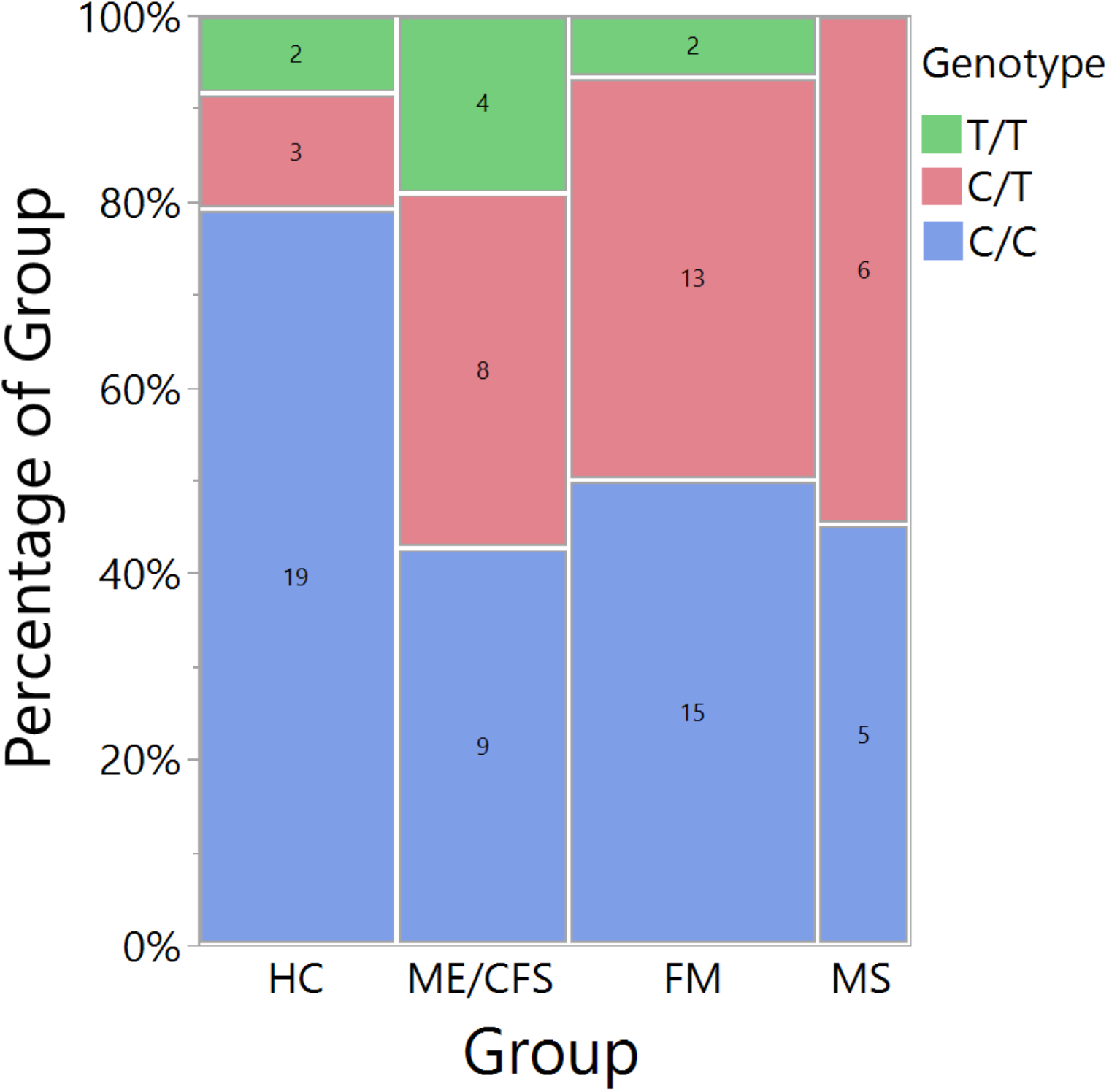
Patient Group by Genotype.

Those with at least one copy of the SNP had significantly higher self-reported fatigue severity (z = 1.99, *p* = 0.046) and higher depression scores (z = 3.03, *p* = 0.0025). Reported pain severity (z = 1.74, *p* = 0.083) and anxiety scores (z = 1.42, *p* = 0.15) were not significantly different between those with and without the polymorphism. Median fatigue and depression scores for each genotype can be seen in Figure 2.

**Fig 2.**
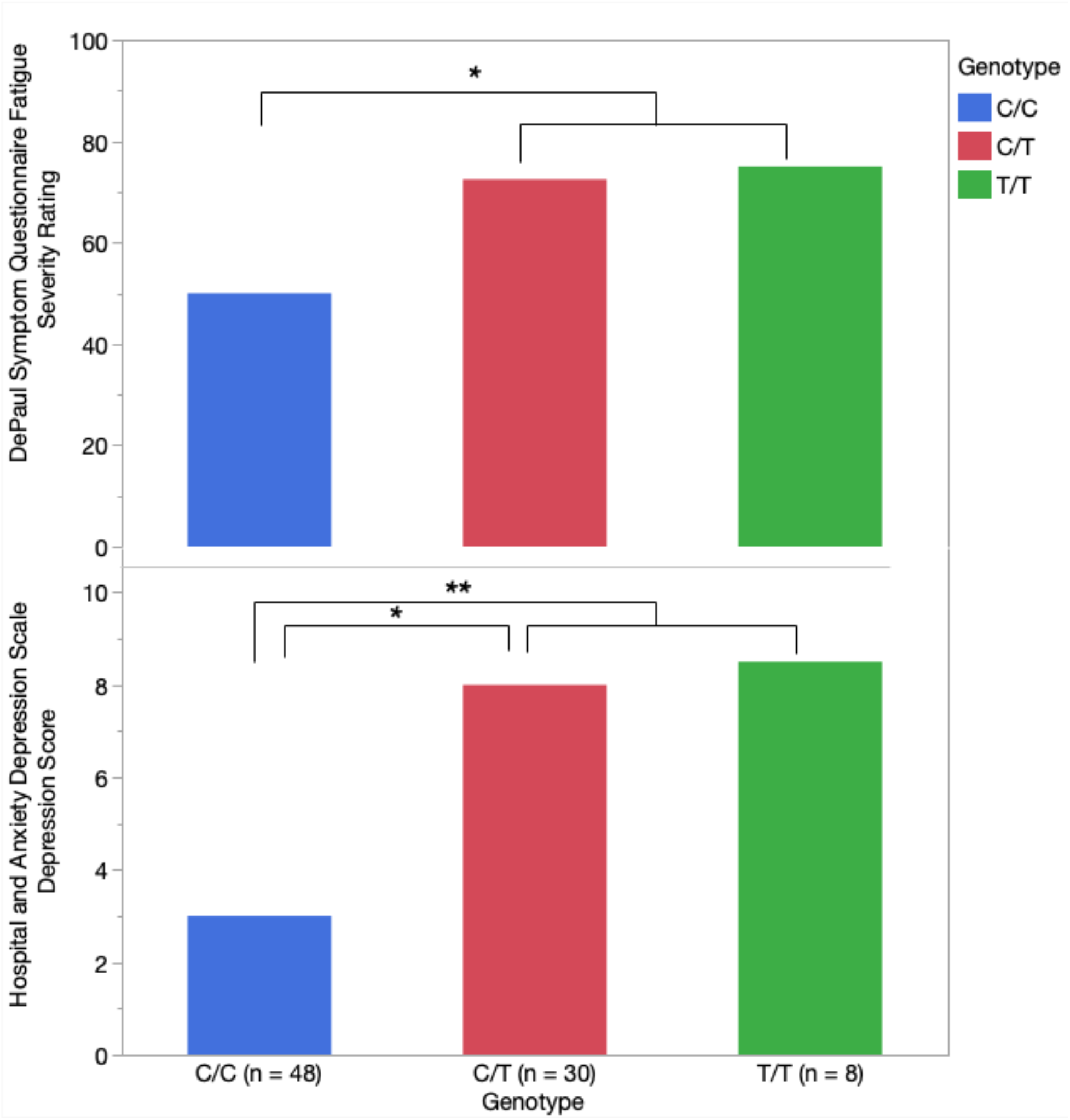
Median Fatigue and Depression Scores. *\*p < 0.05* *\*\*p < 0.01*

When comparing across all three genotypes, only depression scores were significantly different across genotypes (χ ^2^(2) = 9.25, *p* = 0.0098). Pairwise comparisons indicated significantly higher depression scores in the mixed affinity (C/T) group compared to the high affinity group (C/C) (z = 2.77, *p* = 0.016) (Table 1, Figure 2).

**Table 1.** Symptoms by Genotype.

| Symptom Report Means (Standard Deviation) |  |  |  |  |
| --- | --- | --- | --- | --- |
| Genotype | HADS<br>Depression<br>Score<br>(0–21) | HADS<br>Anxiety<br>Score<br>(0–21) | DePaul Fatigue<br>Severity Rating<br>(0–100) | Brief Pain<br>Inventory<br>Severity<br>Score (0–10) |
| C/C | 4.15 (4.30) | 7.46 (4.66) | 44.19 (38.28) | 3.29 (2.94) |
| C/T | 7.38 (5.19) | 9.21 (5.17) | 65.96 (29.68) | 4.55 (2.36) |
| T/T | 7.83 (4.87) | 8.50 (5.24) | 49.29 (46.76) | 3.71 (3.28) |
| Kruskal-Wallis $\chi^2$ | 9.25 | 2.06 | 4.66 | 3.42 |
| *p*-value | **0.0098** | 0.36 | 0.10 | 0.18 |

Note. HADS = Hospital Anxiety and Depression Scale.

## Discussion

To our knowledge, this is the first study to demonstrate a greater frequency of the rs6971 TSPO polymorphism in ME/CFS, FM, and MS, as well as a relationship between the presence of the alternate allele and symptom severity. A prior study investigated the relationship between TSPO genotype, serotonin transporter gene, and fMRI activation during pressure pain in a European FM sample, but did not compare genotypes rates to healthy controls [24]. Our findings suggest that TSPO genotype may be relevant to the pathophysiology of pain and fatigue disorders, though the exact functional consequences of the SNP at rs6971 need further investigation.

TSPO is thought to play a critical role in the early steps of steroidogenesis by transporting cholesterol into mitochondria to be synthesized into pregnenolone. Some evidence suggests that the alanine to threonine substitution at the fifth transmembrane domain reduces pregnenolone production and is associated with higher circulating levels of low-density lipoproteins (LDL) in humans [25]. Evidence has been inconsistent, however, as some TSPO knock-out rodent models have demonstrated intact cholesterol transport and normal pregnenolone levels (for a review see Selvaraj, Stocco [26]). In an effort to resolve these discrepancies, a specific TSPO mutant rodent line lacking the fifth transmembrane domain, corresponding to the rs6971 polymorphism, was developed [27]. These rodents were found to have reduced circulating testosterone, reduced allopregnanolone in the brain, and attenuated cortisol response to ACTH challenge. TSPO genotype may therefore have downstream effects on hypothalamic-pituitary-adrenal (HPA) axis regulation. In addition to HPA axis function, steroidogenesis by TSPO may also play a direct role in immune function by modulating cytokine release [28]. Currently, however, there is no evidence of altered immune processes associated with TSPO genotype in humans, however TSPO agonists have shown beneficial effects on inflammation regulation in preclinical studies [29-32].

The rs6971 SNP has been primary investigated in psychiatric illness due to its potential influence on HPA axis regulation. The SNP has been associated with higher rates of bipolar disorder [33], variance in diurnal cortisol rhythms in bipolar disorder [34], and higher cholesterol, low density lipoprotein (LDL), triglycerides, and withdrawal symptoms in alcohol use disorder [35]. Presence of the polymorphism has also been associated with separation anxiety [36], and panic disorder [37]. In addition to psychiatric illness, the SNP has been associated with poorer prognosis in male glioblastoma patients [38] and is possibly related to Parkison’s disease progression [39].

Alterations in HPA axis function, specifically hypocortisolism and hyporesponsiveness to ACTH challenge, have consistently been observed in ME/CFS patients (for a review see Morris, Anderson [40]). Similar, but more variable results have been found in FM (for a meta-analysis see Beiner, Lucas [41]). Additionally, lower pregnenolone or pregnenolone-steroid ratios have been seen in women with ME/CFS [42] and FM [43]. Further investigation is warranted on how the presence of the rs6971 polymorphism potentially may contribute to these neuroendocrine abnormalities.

### Limitations

There are several limitations to the present study. This exploratory analysis arose from incidental observations during the course of a parent study and requires replication in larger, more diverse samples of both men and women. The parent study also did not include the collection of relevant hormones or lipids, limiting the ability of the study to test the proposed consequences of TSPO genotype.

The TSPO SNP is also known to vary by ancestry; East-Asian populations show the lowest rates of the alternate allele polymorphism (96.9% high affinity binders), followed by African populations (67.4% high affinity binders), American populations (69.2% high affinity binders) and the greatest rates of the polymorphism is found in European populations (45.7% high affinity binders) [44]. In our United States based sample, 79% of healthy controls were high affinity binders, which may be higher than general American population rates due to our exclusion of medical conditions and significant pain, fatigue, or mental health conditions. Although the present study compared genotypes across racial/ethnic groups, future studies should include more diverse and balanced samples, and collect more detail on ethnic background. Ethnicity was also self-reported by participants who may not know or fully disclose their ethnic makeup.

### Future Directions

If these findings are confirmed in larger and diverse samples, future investigations should incorporate direct measures of neurosteroids, HPA axis function, and immune markers. Additional avenues of study may be the potential benefit of TSPO agonism by ligands to increase neurosteroid production or modulate neuroinflammatory processes. TSPO drug ligands have been trialed in anxiety (e.g., Etioxine, Emapunil), and recently in rodent models of Alzheimer’s disease [31] and multiple sclerosis [32]. These have yet to be formally investigated in FM or ME/CFS. The efficacy of such treatments, however, is likely to vary by rs6971 genotype [45], highlighting the importance of testing and considering genetic variation in the development of TSPO based treatments.

## Data Availability

All data produced in the present study are available upon reasonable request to the authors

## Acknowledgements

We thank the participants for their time and contribution. We thank the UAB Specimen Processing and Biorepository Unit (SPAN) and Genomic Science Core Laboratories for their services.

## Authorship Declaration

All authors have made significant contributions to the work and are in full agreement with the content of the manuscript. This analysis was designed by C.J. and J.Y. The data were analyzed by C.J. and the results were critically examined by all authors. C.J. had a primary role in preparing the manuscript which was edited by J.Y. All authors have approved the final version of the manuscript.

